# Operationalization of Severe Maternal Morbidity surveillance in administrative databases of the Brazilian public and private health systems: Brazil 2015-2022

**DOI:** 10.1101/2025.08.20.25334044

**Authors:** Rosa Maria Soares Madeira Domingues, Marcos Augusto Bastos Dias, Luís Carlos Torres Guillen, Rejane Sobrino Pinheiro, Valeria Saraceni, Claudia Medina Coeli

## Abstract

The study of severe maternal morbidity (SMM) is an important strategy for improving the quality of obstetric care. The objective of this study is to assess SMM cases recorded in the Hospital Information System of the Unified Health System (SIH/SUS), which contains public funded hospitalizations, and in the database of the National Supplementary Health Agency (ANS), which contains hospitalizations paid for by health plans. This is a cross-sectional study using obstetric hospitalizations of women aged 10 to 49 years from both databases in the period 2015 to 2022. We assessed SMM cases using the World Health Organization’s criteria for potentially life-threatening conditions (PLTC). Of the 26 PLTC criteria, 22 were operationalized through diagnoses during hospitalization, using International Classification of Diseases-10 codes, or medical procedures, using specific codes of each database. We estimated the frequency and odds ratio of death for each isolated criterion, for grouped criteria and for the total number of SMM cases, as well as the frequency of deaths according to the number of SMM criteria. We identified 18,807,757 obstetric hospitalizations in the SIH/SUS and 3,776,986 in the ANS database. The estimated proportion of SMM was 8.51% and 7.30% in SIH/SUS and ANS, respectively. Most cases presented only one criterion. Differences were observed among sectors, with a higher frequency of procedure-based criteria in the ANS database, compatible with the greater availability of resources in private services. The proportion of deaths increased with the increase in the number of criteria, and more severe criteria presented a higher odds ratio of death. The results indicate that it is possible to estimate the number of SMM cases in the country, both in public and private hospitalizations. Continued investment in the quality of recording in both databases are necessary so that the estimates can truly reflect the maternal morbidity of Brazilian women.

## Introduction

Maternal death is a serious public health problem that disproportionately affects less developed countries [1]. Within the scope of the Sustainable Development Goals (SDG), there is a target of reducing the global maternal mortality ratio (MMR) to 70 cases per 100,000 live births (LB) by 2030, with this target being 30 per 100,000 LB for Brazil [2].

In 2023, the global MMR was estimated at 197 deaths per 100,000 live births, with an annual downward trend of 2.2% between 2000 and 2023, insufficient to reach the 2030 target [1]. In Brazil, the MMR also showed a downward trend between 2000 and 2023, remaining close to 50 deaths per 100,000 live births. During the COVID-19 pandemic, a significant increase in the MMR was observed in the country, reaching 113.14 deaths per 100,000 live births in 2021, returning to pre-pandemic levels in 2022 and 2023 [3].

Despite the still high rates in Brazil, maternal deaths are a low-frequency event, concentrated in larger municipalities. Between 2012 and 2020, 2,389 Brazilian municipalities (approximately 43%) did not record maternal deaths [4], making it difficult to develop strategies to improve the quality of maternal health care based on such an infrequent event.

Since 2009, the World Health Organization (WHO) has recommended analyzing severe maternal morbidity as a complementary strategy to maternal death surveillance, as these events are more frequent and share the same determinants as maternal death, allowing for more robust analyses [5]. Within the spectrum of severe maternal morbidity, cases of life-threatening conditions, also called maternal near miss (MNM), are at the extreme end of the severity spectrum, along with maternal deaths, and are characterized by clinical, laboratory, and management criteria that indicate organ dysfunction. Cases of potentially life-threatening conditions include other serious conditions, but not characterized solely by organ dysfunction, and are classified according to 26 criteria, including hypertensive disorders, bleeding disorders, other systemic disorders, and indicators of management severity [6].

In Brazil, previous studies have assessed severe maternal morbidity using either hospital record data [7,8] or the Hospital Information System of the Unified Health System (SIH-SUS) [9–13], a system implemented in 1990 that aims to pay for publicly funded hospitalizations in public or private hospitals affiliated with the SUS. These studies used both the criteria proposed by the WHO and previous criteria based on the Mandel or Waterstone classifications [11,12]. It is noteworthy that no studies were identified that assessed maternal morbidity in privately funded hospitalizations, whose databases have been available since 2015.

In studies using data from the SIH/SUS [9–13], the different criteria used to operationalize the definition of severe maternal morbidity make it difficult to compare results. One study evaluated the validity of the SIH/SUS for studying MNM cases according to the WHO classification, finding low sensitivity and positive predictive value [10]. This result can be explained by the difficulty in operationalizing this definition with the data available in the SIH/SUS, as these criteria use laboratory test results, clinical parameters, and duration of symptoms and/or interventions, which are not available in the SIH/SUS. This leads to the use of approximate criteria based on diagnoses and procedures, resulting in case classification errors. Potentially life-threatening conditions (PLTC) are based on clinical diagnoses and procedures closer to the diagnostic and procedural information available in the SIH/SUS and the National Supplementary Health Agency (ANS) database.

Considering the importance of studying severe maternal morbidity for reducing maternal death and the availability of national administrative databases containing morbidity data, this study aims to present a methodological proposal for assessing cases of maternal severe morbidity recorded in the administrative databases of SIH/SUS and ANS using the definition of a PLTC proposed by the WHO [6].

## Materials and methods

### Study design

This is a cross-sectional study using de-identified, publicly accessible databases from the SIH/SUS and the ANS hospital admissions database, both for the period 2015-2022.

### Study Population

Women aged 10 to 49 with obstetric hospitalizations in the SIH/SUS and the supplementary health databases.

### Data Source

Data from the SIH/SUS were captured by the microdatasus package using the R statistical programming language [14] (https://github.com/rfsaldanha/microdatasus). Data from the

ANS databases were obtained from https://dados.gov.br/dados/conjuntos-dados/procedimentos-hospitalares-por-uf. Both databases were accessed on December 2, 2024.

The SIH/SUS provides two databases: the “reduced database,” which contains admission diagnoses according to the International Classification of Diseases (ICD) and the main procedure performed; and the “professional services database,” which contains all professional acts performed during the hospitalization. In the reduced database, admission diagnoses are recorded in several fields (principal diagnosis, secondary diagnoses 1 to 9, ICD_notification, ICD_death, and ICD_associated). The two databases can be linked using the AIH number. The AIH (Hospitalization Authorization, from the Portuguese acronym for *Autorização de Internação Hospitalar*) is the form filled for each discharge based on the medical record abstract.

The ANS database also provides two databases: the “consolidated database,” which contains the hospitalization ICD available in four fields (ICD_1, ICD_2, ICD-3, and ICD_4), and the “detailed database,” which contains all procedures performed during hospitalization. The two databases can be linked by the variable ID_EVENTO_ATENCAO_SAUDE.

The diagnoses recorded in both databases are those available in the International Classification of Diseases version 10 (ICD-10). The procedures used in SIH/SUS follow the SIGTAP table (SUS Procedure, Medication and OPM Table Management System), available at http://sigtap.datasus.gov.br/tabela-unificada/app/sec/inicio.jsp), while the procedures used in the ANS database follow the Unified Terminology of Supplementary Health (TUSS) available at https://dados.gov.br/dados/conjuntos-dados/terminologia-unificada-da-saude-suplementar-tuss.

### Identification of Obstetric Admissions

In the SIH/SUS, there is no specific field for identifying obstetric admissions. The operational definitions described in Table 1, based on ICD and procedures, were used to select obstetric admissions among hospital admissions of women aged 10 to 49. However, before selecting these hospitalizations, it was necessary to identify the episode of care. In the SIH/SUS, in some specific situations, such as when a surgical procedure is indicated during an initial clinical admission or vice versa, a single hospitalization episode may contain more than one AIH. An algorithm was then developed to identify episodes of care with more than one AIH [15]. Compared to episodes with only one AIH, episodes with multiple AIH had longer hospital stays, higher costs, and a higher proportion of complications and deaths [15]. After identifying the episode of care, any episode of care, with one or multiple AIH, containing an ICD or procedure described in the criteria in Table 1, was classified as an obstetric hospitalization.

**Table 1.** Criteria used to identify obstetric hospitalizations in the Hospital Information System of the Unified Health System (SIH-SUS) and in the National Supplementary Health Agency (ANS) database.

| Criteria/database | SIH/SUS | ANS |
| --- | --- | --- |
| Type of hospitalization | Information not available | Obstetric hospitalization (variable “type of hospitalization=3) except if the admission diagnosis is different from P0, P1, P2, P3, P4, P5, P20, P35, P37, P50, P56, P93, P95, P96.<br>**AND/OR** |
| Diagnosis at hospital admission | ICD of group “O” registered in any of the diagnosis fields (principal diagnosis, secondary diagnoses 1 to 9, ICD_notification, ICD_death, ICD_associated) | ICD of group “O or ICD “Z34 to Z39” or the following ICD of group “P” (P000-P036, P038-P046, P048, P049, P051, P052, P059, P100- P104, P108- P115, P119-P201, P209-, P211, P219, P350 - P353, P358, P359, P370, P371, P373, P374, P378, P379, P392, P398, P399, P500- P505, P508, P509, P520- P526, P528, P529, P53, P550, P551, P558, P559, P560, P569, P60, P832, P833, P93, P95, P964, |
|  |  | P965, P968, P969) if the type of hospitalization is not pediatric, recorded in the ICD_1, ICD_2, ICD_3 or ICD_4 fields |
|  | **AND/OR** | **AND/OR** |
| Procedures | Any of the following procedures:<br>02.01.01.001-1, 02.11.04.001-0,<br>02.11.04.006-1, 03.10.01.001-2,<br>03.10.01.002-0, 03.10.01.003-9,<br>03.10.01.004-7, 03.10.01.005-5,<br>03.03.10.001-0, 03.03.10.002-8,<br>03.03.10.003-6, 03.03.10.004-4,<br>03.03.10.005-2, 04.09.06.001-1,<br>04.09.06.005-4, 04.09.06.007-0,<br>04.11.01.001-8, 04.11.01.002-6,<br>04.11.01.003-4, 04.11.01.004-2,<br>04.11.01.005-0, 04.11.01.007-7,<br>04.11.01.008-5, 04.11.02.001-3,<br>04.11.02.002-1, 04.11.02.003-0,<br>04.11.02.004-8, 04.17.01.002-8,<br>04.17.01.001-0, 04.17.01.003-6 | Any of the following procedures:<br>31309062, 31303013, 31309020, 40501124,<br>40501132, 40501221, 40503097, 20202016,<br>20202024, 31309011, 31309038, 31309097,<br>31309127, 31309135, 31309054, 31309208,<br>31309186, 31309089, 31309100, 31309119,<br>31309194, 31309151, 31303323, 31303285,<br>31303315, 31306047, 31309178, 31309046,<br>31309216, 31309224, 31309232, 40201015,<br>31309259, 40309444, 41401670, 40502139,<br>40502147, 40502155, 40901238, 40901246,<br>40901254, 40901262, 40901270, 40901289,<br>40901297, 40902013, 40902021, 40901505,<br>40901556 |

The ANS database contains a field for recording obstetric admissions. However, the comparison of this information with the diagnoses and procedures performed during the hospitalization showed that using this information alone would result in both underreporting of obstetric admissions (primarily due to underreporting of admissions for abortion, but also admissions for cesarean sections) and overreporting due to the inclusion of neonatal admissions, since admissions of newborns in the first days of life are generally recorded in the insured’s hospitalization (data not shown in the table). Therefore, to identify obstetric admissions in the ANS database, we used the criteria described in Table 1. It should be noted that, by court order, ICD registration is not mandatory for supplementary healthcare admissions, with the national proportion of ignored data being 30%, with significant variation by state (data not shown in the table).

### Identification of Severe Maternal Morbidity (SMM) cases

We used the 26 criteria proposed by the WHO to identify PLTC [6], which will be referred to in this study as SMM. Cases were classified according to the ICD in both the SIH/SUS and the ANS databases, while the classification by procedure followed the items available in SIGTAP and TUSS, for hospitalizations in the SIH/SUS and the ANS databases, respectively. The criteria used in both databases are described in Table 2.

**Table 2.**
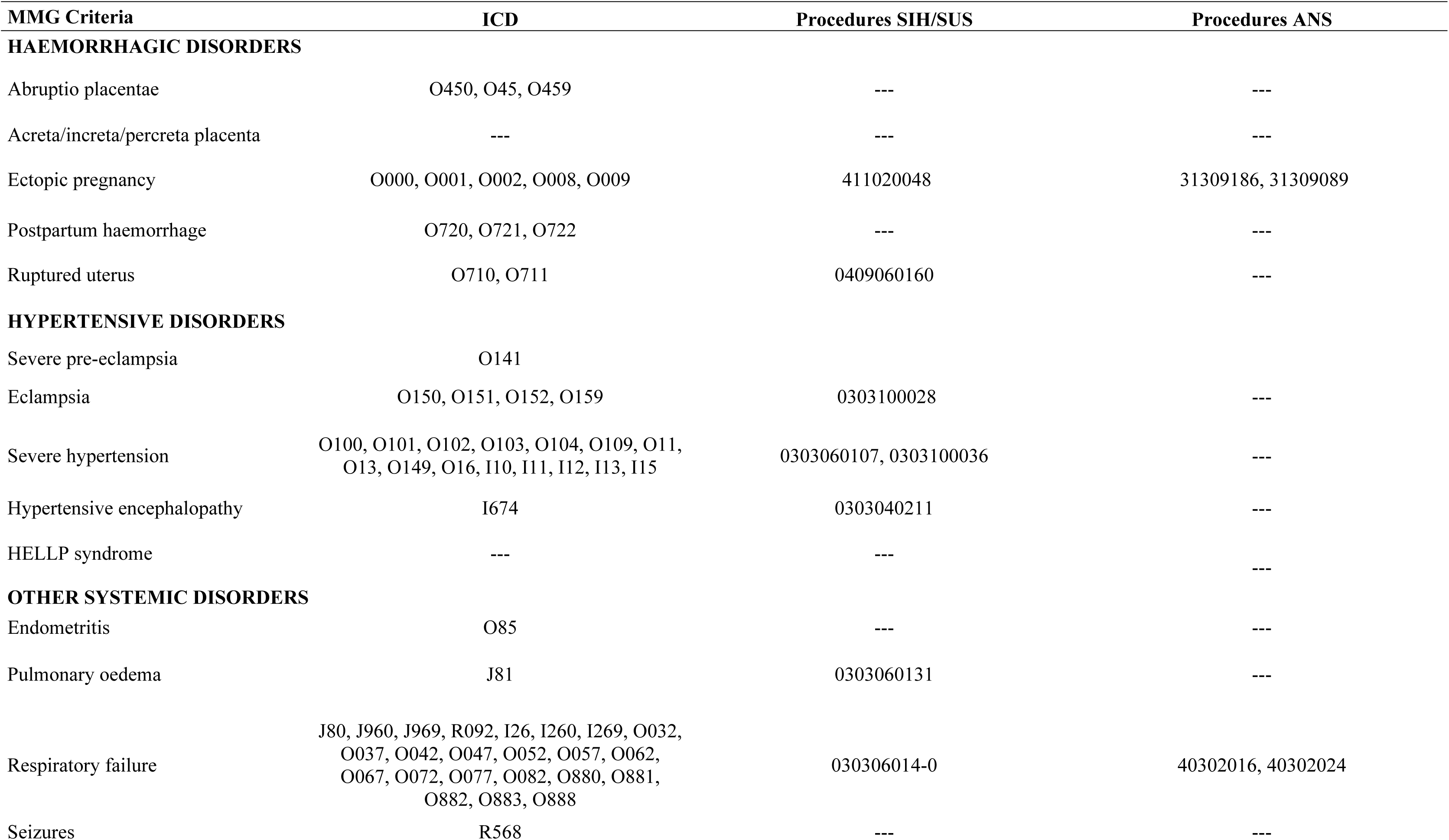
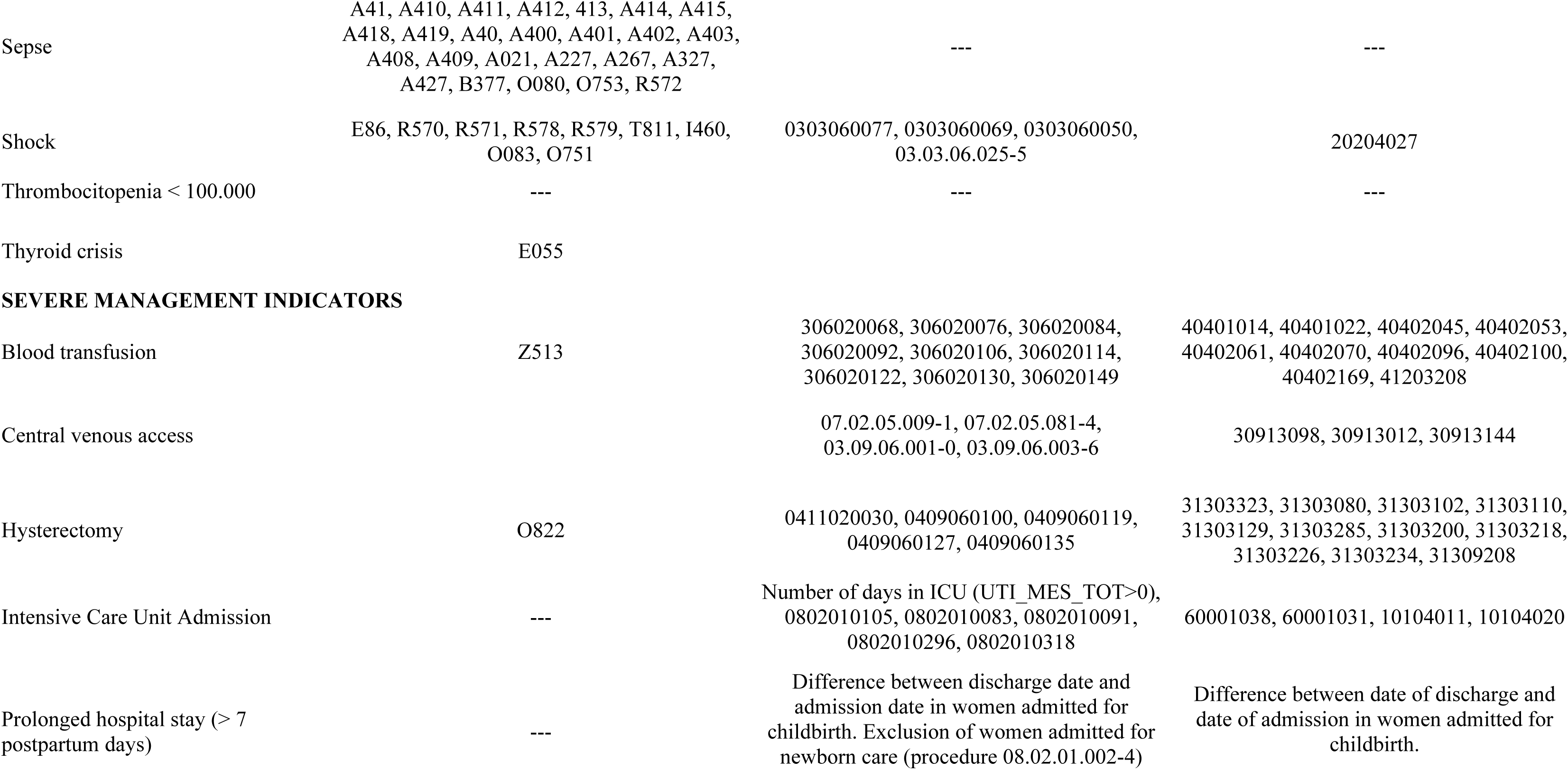
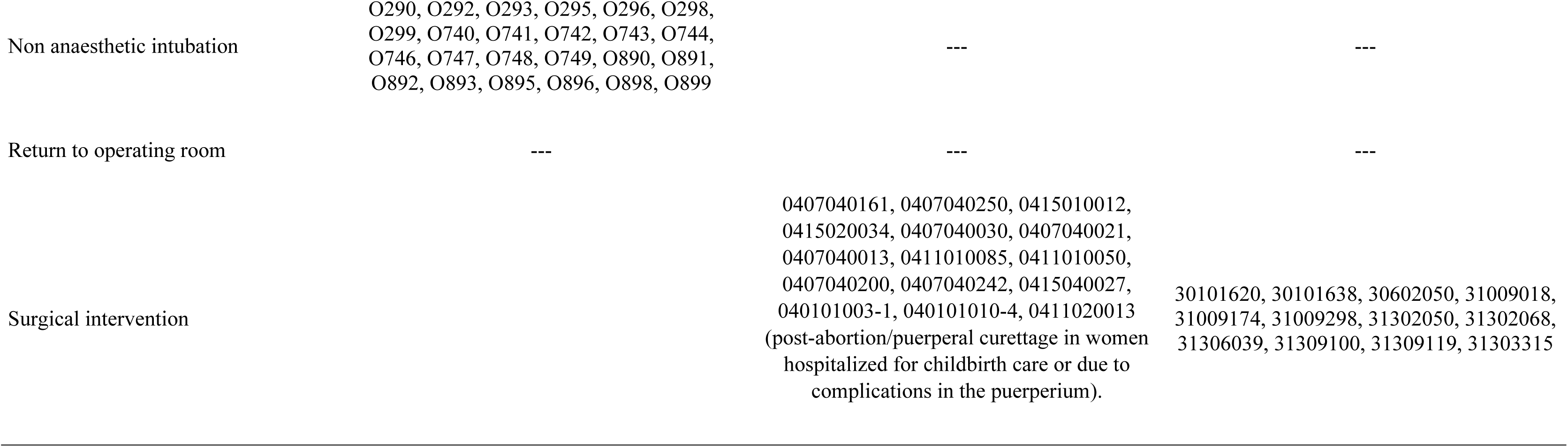
Criteria used to diagnose cases of Severe Maternal Morbidity (SMM) in the Hospital Information System of the Unified Health System (SIH/SUS) and the National Supplementary Health Agency database (ANS).

### Data Analysis

In both databases, we verified the frequency of each criterion in relation to the total number of obstetric admissions and the total number of SMM cases. We also analyzed the frequency of cases according to the number of criteria met.

To assess construct validity (convergent validity) [16], we evaluated the association between SMM (globally, by grouped criteria, and by isolated criterion) and death using odds ratios. For this analysis, we only included admissions that resulted in discharge alive or in-hospital death. The exclusion of admissions that resulted in hospital stay, inter-hospital transfer, or unreported discharge aimed to analyze only admissions with a known outcome regarding the woman’s survival. We estimated the odds ratio for in-hospital death using all episodes of care that did not meet the criterion under analysis as the reference. Hence, the reference included episodes that met other criteria or did not meet any SMM criteria. Furthermore, we verified the frequency of deaths during hospitalization for the total number of obstetric hospitalizations and according to the number of SMM criteria. R software [14] was used in all analyses.

### Ethical Aspects

We only used de-identified publicly available data. According to The Brazilian National Health Council Ethics Resolution n*◦* 510/2016 (April 7, 2016) the research ethics committee approval is waived.

## Results

From 2015 to 2022, 18,807,757 obstetric hospitalizations were identified in the SIH/SUS database and 3,776,986 in the ANS database, with 8.51% (1,600,796/18,807,757) and 7.30% (275,731/3,776,986) of hospitalizations classified as SMM, respectively.

The most frequent SMM criterion in SIH/SUS was severe hypertension (40.75%), followed by prolonged hospital stay (PLS, postpartum length of stay greater than seven days) (18.28%), severe pre-eclampsia (12.38%) and blood transfusion (11.63%), while in ANS the most frequent criteria were Intensive Care Unit (ICU) admission (34.67%), PLS (20.67%), severe hypertension (18.46%) and respiratory failure (12.94%). Considering the grouped criteria [6], the highest frequency in the ANS database was severe management indicators (62.66%), followed by hypertensive disorders (22.19%), other systemic disorders (14.08%) and haemorrhagic disorders (12.47%), while in the SIH/SUS database the most frequent grouped criteria were hypertensive disorders (54.06%), severe management indicators (39.77%), haemorrhagic disorders (13.62%) and other systemic disorders (4.28%).

When comparing the proportion of each criterion in relation to the total number of obstetric hospitalizations in each database, it was observed that the criteria respiratory failure, central venous access, ICU admission and hysterectomy were more frequent in the ANS than in the SIH/SUS, with values 17.0, 7.5, 4.7 and 4.2 higher, respectively, while ruptured uterus, eclampsia and severe pre-eclampsia, abruptio placentae and endometritis presented values 4 to 7 times higher in the SIH/SUS, when compared to the ANS (Table 3).

**Table 3:** Frequency of Severe Maternal Morbidity (SMM) cases in the Hospital Information System of the Unified Health System (SIH-SUS) and the National Supplementary Health Agency (ANS) database. Brazil, 2015-2022.

| CRITERIA SMM | Obstetric Hospitalizations SIH/SUS<br>(N=18,807,757) |  |  | Obstetric Hospitalizations ANS<br>(N=3,776,986) |  |  | RATIO SIH/ANS |  |
| --- | --- | --- | --- | --- | --- | --- | --- | --- |
|  | SMM cases | % Obstetric Hospitalizations | % SMM cases | SMM cases | % Obstetric Hospitalizations | % SMM cases | Criteria/total hospitalizations | Criteria/total SMM cases |
| **HAEMORRHAGIC DISORDERS** |  |  |  |  |  |  |  |  |
| Abruptio placentae | 61,056 | 0.32 | 3.81 | 2,839 | 0.08 | 1.03 | 4.32 | 3.70 |
| Ectopic pregnancy | 130,436 | 0.69 | 8.15 | 30,112 | 0.80 | 10.92 | 0.87 | 0.75 |
| Postpartum haemorrhage | 22,546 | 0.12 | 1.41 | 1,302 | 0.03 | 0.47 | 3.48 | 2.98 |
| Ruptured uterus | 4,287 | 0.02 | 0.27 | 122 | 0.00 | 0.04 | 7.06 | 6.05 |
| **TOTAL HAEMORRHAGIC DISORDERS** | **218,017** | **1.16** | **13.62** | **34,374** | **0.91** | **12.47** | **1.27** | **1.09** |
| **HYPERTENSIVE DISORDERS** |  |  |  |  |  |  |  |  |
| Severe pre-eclampsia | 198,247 | 1.05 | 12.38 | 8,183 | 0.22 | 2.97 | 4.87 | 4.17 |
| Eclampsia | 56,430 | 0.30 | 3.53 | 2,233 | 0.06 | 0.81 | 5.07 | 4.35 |
| Severe hypertension | 652,396 | 3.47 | 40.75 | 50,899 | 1.35 | 18.46 | 2.57 | 2.21 |
| Hypertensive encephalopathy | 20 | 0.00 | 0.00 | 3 | 0.00 | 0.00 | 1.34 | 1.15 |
| **TOTAL HYPERTENSIVE DISORDERS** | **865,446** | **4.60** | **54.06** | **61,181** | **1.62** | **22.19** | **2.84** | **2.44** |
| **OTHER SYSTEMIC DISORDERS** |  |  |  |  |  |  |  |  |
| Endometritis | 51966 | 0.28 | 3.25 | 2,093 | 0.06 | 0.76 | 4.99 | 4.28 |
| Pulmonary oedema | 208 | 0.00 | 0.01 | 23 | 0.00 | 0.01 | 1.82 | 1.56 |
| Respiratory failure | 10,485 | 0.06 | 0.65 | 35,681 | 0.94 | 12.94 | 0.06 | 0.05 |
| Seizures | 627 | 0.00 | 0.04 | 107 | 0.00 | 0.04 | 1.18 | 1.01 |
| Sepsis | 4,218 | 0.02 | 0.26 | 698 | 0.02 | 0.25 | 1.21 | 1.04 |
| Shock | 1,385 | 0.01 | 0.09 | 379 | 0.01 | 0.14 | 0.73 | 0.63 |
| Thyroid crisis | 12 | 0.00 | 0.00 | 4 | 0.00 | 0.00 | 0.60 | 0.52 |
| **TOTAL OTHER SYSTEMIC DISORDERS** | **68,514** | **0.36** | **4.28** | **38,836** | **1.03** | **14.08** | **0.35** | **0.30** |
| **SEVERE MANAGEMENT INDICATORS** |  |  |  |  |  |  |  |  |
| Blood transfusion | 186,101 | 0.99 | 11.63 | 12,806 | 0.34 | 4.64 | 2.92 | 2.50 |
| Central venous access | 2,967 | 0.02 | 0.19 | 4,482 | 0.12 | 1.63 | 0.13 | 0.11 |
| Hysterectomy | 12,523 | 0.07 | 0.78 | 10,670 | 0.28 | 3.87 | 0.24 | 0.20 |
| ICU admission | 100,585 | 0.53 | 6.28 | 95,596 | 2.53 | 34.67 | 0.21 | 0.18 |
| Prolonged hospital stay (> 7 postpartum days) | 292,625 | 1.56 | 18.28 | 56,980 | 1.51 | 20.67 | 1.03 | 0.88 |
| Non anaesthetic intubation | 2,734 | 0.01 | 0.17 | 308 | 0.01 | 0.11 | 1.78 | 1.53 |
| Surgical intervention | 131,494 | 0.70 | 8.21 | 17,911 | 0.47 | 6.50 | 1.47 | 1.26 |
| **TOTAL SEVERE MANAGEMENT INDICATORS** | **636,559** | **3.38** | **39.77** | **172,780** | **4.57** | **62.66** | **0.74** | **0.63** |
| **TOTAL SMM** | **1,600,796** | **8.51** |  | **275,731** | **7.30** |  | **1.17** | **-** |

In the SIH/SUS database, 99% (18,628,749/18,807,757) of obstetric hospitalizations had a known outcome (hospital discharge or death), while in the ANS database this value was 94.8% (3,580,965/3,776,986). A greater chance of death was observed in criteria of greater severity, mainly in the SIH/SUS, with the highest values observed in the criteria shock (OR = 692.78), central venous access (OR = 517.17), pulmonary oedema (OR = 438.70) and hypertensive encephalopathy (OR = 356.88), while in the ANS database the highest odds ratios were observed in the criteria shock (OR = 52.52), central venous access (OR 45.80), seizures (OR = 18.19), blood transfusion (OR = 15.99) and ruptured uterus (OR = 15.08). Hospitalizations in the SIH/SUS presented a higher odds ratio of death in all criteria, with the greatest differences observed in the criteria hysterectomy, postpartum haemorrhage, sepse, shock, ICU admission and central venous access (Table 4).

**Table 4:** Odds ratio of death according to Severe Maternal Morbidity criteria in the Hospital Information System of the Unified Health System (SIH-SUS) and the National Supplementary Health Agency (ANS) database. Brazil, 2015-2022.

| CRITERION | Hospitalizations SIH/SUS*<br>(N=18.628.749) |  |  | Hospitalizations ANS*<br>(N= 3.545.882) |  |  | SIH/ANS<br>OR<br>RATIO |
| --- | --- | --- | --- | --- | --- | --- | --- |
|  | SMM<br>death | SMM<br>alive | OR | SMM<br>death | SMM<br>alive | OR |  |
| HAEMORRHAGIC DISORDERS |  |  |  |  |  |  |  |
| Abruptio placentae | 160 | 59,899 | 6.80 | 15 | 2,634 | 4.78 | 1.42 |
| Ectopic pregnancy | 85 | 128,931 | 1.65 | 8 | 28,847 | 0.23 | 7.17 |
| Postpartum haemorrhage | 240 | 21,096 | 29.32 | 2 | 1,243 | 1.35 | 21.72 |
| Ruptured uterus | 34 | 4,104 | 20.78 | 2 | 111 | 15.08 | 1.38 |
| Total haemorrhagic disorders | 504 | 213,774 | 6.24 | 27 | 32,834 | 0.69 | 9.04 |
| HYPERTENSIVE DISORDERS |  |  |  |  |  |  |  |
| Severe pre-eclampsia | 320 | 190,572 | 4.34 | 6 | 7,408 | 0.68 | 6.38 |
| Eclampsia | 330 | 52,443 | 16.40 | 12 | 1,967 | 5.11 | 3.21 |
| Severe hypertension | 544 | 636,331 | 2.22 | 59 | 47,539 | 1.04 | 2.13 |
| Hypertensive encephalopathy | 2 | 14 | 356,88 | 0 | 3 | - | - |
| Total hypertensive disorders | 1,070 | 839,754 | 3.55 | 77 | 56,792 | 1.14 | 3.11 |
| OTHER SYSTEMIC DISORDERS |  |  |  |  |  |  |  |
| Endometritis | 162 | 50,169 | 8.22 | 8 | 1,949 | 3.44 | 2.39 |
| Pulmonary oedema | 28 | 160 | 438.70 | 0 | 17 | - | - |
| Respiratory failure | 217 | 10,110 | 55.18 | 282 | 30,955 | 8.09 | 6.82 |
| Seizures | 9 | 586 | 38.40 | 2 | 92 | 18.19 | 2.11 |
| Sepsis | 219 | 3,819 | 147.52 | 8 | 602 | 11.13 | 13.25 |
| Shock | 269 | 1,006 | 692.78 | 20 | 320 | 52.52 | 13.19 |
| Thyroid crisis | 0 | 12 | - | 0 | 4 | 0.00 | - |
| **Total other systemic disorders** | 785 | 65,630 | 33.27 | 313 | 33,818 | 8.27 | 4.02 |
| **SEVERE MANAGEMENT INDICATORS** |  |  |  |  |  |  |  |
| Blood transfusion | 2,181 | 175,881 | 43.36 | 206 | 11,287 | 15.99 | 2.71 |
| Central venous access | 395 | 2,014 | 517.17 | 183 | 3,488 | 45.80 | 11.29 |
| Hysterectomy | 513 | 10,660 | 129.00 | 56 | 10,037 | 4.72 | 27.33 |
| ICU admission | 2,579 | 89,783 | 109.15 | 769 | 84,250 | 9.09 | 12.01 |
| Prolonged hospital stay (> 7 postpartum days) | 568 | 288,306 | 5.24 | 234 | 46,139 | 4.43 | 1.18 |
| Non anaesthetic intubation | 11 | 2,654 | 10.37 | 1 | 291 | 2.87 | 3.61 |
| Surgical intervention | 453 | 127,123 | 9.41 | 68 | 16,772 | 3.43 | 2.74 |
| **Total severe management indicators** | 3,556 | 614,525 | 26.72 | 914 | 150,975 | 6.17 | 4.33 |
| **TOTAL SMM** | 4,104 | 1,554,823 | 13.44 | 1,016 | 246,858 | 4.15 | 3.24 |

\*Obstetric admissions with known outcome (hospital discharge or in-hospital death).
ICU=Intensive Care Unit

In both databases, the largest proportion of cases had only one SMM criterion (84.51% in SIH/SUS [1,317,390/1,558,927] and 85.83% [184,219/214,622]) in ANS. In SIH/SUS, some women presented up to 10 criteria, while in ANS the maximum number was seven criteria (Table 3). The in-hospital mortality rate among cases with a known outcome was 0.04% in SIH/SUS (7,456/18,828,749) and 0.12 (4,276/3,580,965) in the ANS. In both databases, the proportion of deaths increased with the presence of a greater number of SMM criteria, ranging from 0.02% (no criteria) to 100% (10 criteria) in the SIH/SUS and from 0.10% (no criteria) to 8,75% (five criteria) in the ANS. No deaths were observed in women with 6 or 7 criteria in the ANS database, but the number of women in this category was very small. For women with up to four MMG criteria, a higher frequency of deaths was observed in obstetric hospitalizations in the ANS, while a higher proportion of deaths in women with five or more criteria was observed in the SIH/SUS (Table 5).

**Table 5.**
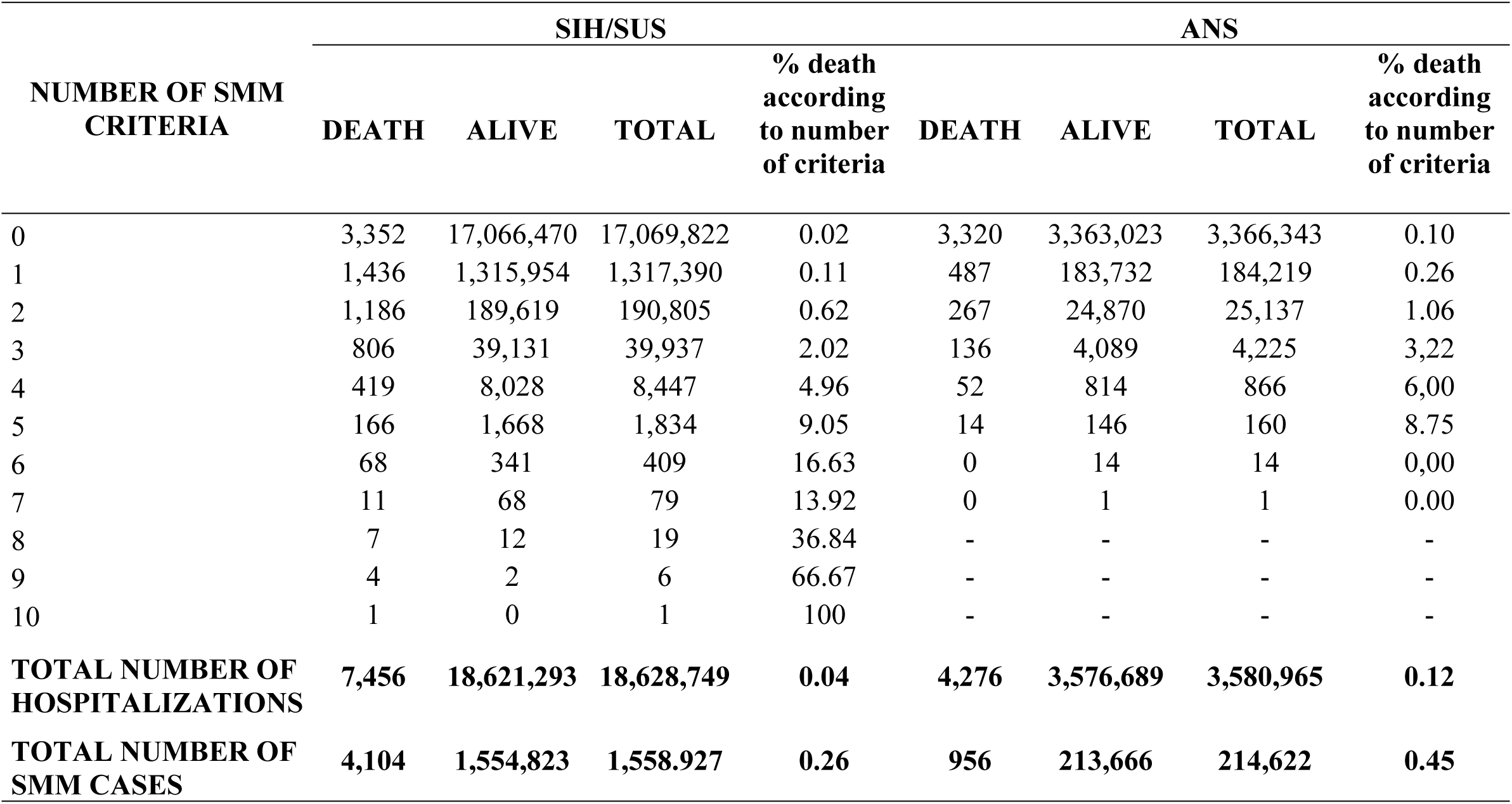
Frequency of deaths according to the number of Severe Maternal Morbidity (SMM) criteria in the Hospital Information System of the Unified Health System (SIH-SUS) and the National Supplementary Health Agency (ANS) database. Brazil, 2015-2022.

## Discussion

The results of this study show that it is possible to estimate SMM using the hospital admissions databases from SIH/SUS and ANS. The estimated proportion of SMM for the 2015-2022 period was 8.51% and 7.30% in SIH/SUS and ANS, respectively. Most cases presented only one criterion. The proportion of deaths increased with the increase in the number of criteria, and more severe criteria presented a higher odds ratio of death.

Only four of the 26 criteria recommended by the WHO for identifying PLTC could not be operationalized with the information available in the two databases: accreta/increta/percreta placenta; HELLP syndrome; thrombocytopenia <100,000; and return to operating room. It should be noted, however, that many cases of “return to operating room” may have been captured using the “surgical procedures” criterion, many of them related to complications requiring return to the operating room. Furthermore, the International Classification of Diseases version 11 (ICD 11) already has a code for HELLP syndrome (JA24.2), which will allow the identification of this condition when ICD 11 is implemented in the country. This result differs from the operationalization of the criteria for identifying MNM cases proposed by the WHO. Of the 25 criteria, only one (cardiopulmonary resuscitation) can be directly operationalized using available procedures. The others require the use of approximate diagnoses and procedures to identify situations of organ dysfunction, resulting in low sensitivity and low predictive value of available administrative data for identifying MNM cases [10]. Difficulties in operationalizing MNM criteria have also been reported in other countries, particularly low-income countries, with some criteria being adapted [17]. The main adaptations identified were the exclusion of laboratory and intervention criteria due to resource limitations, with the number of blood transfusions being the most frequently modified intervention [17].

In a systematic review of 60 studies conducted mostly in low-and middle-income countries, the global frequency of MNM cases was estimated at 1.4% (95% CI 0.4%-2.5%) [17]. Global estimates for PLTC using the criteria proposed by the WHO were not found. Systematic reviews evaluating SMM in low-[18] and high-income countries [18,19] found several definitions in use, with a variable number of criteria based on diagnoses and procedures. A higher frequency of SMM was observed in low-income countries, mainly in sub-Saharan Africa. In all countries, the main causes of SMM were severe obstetric hemorrhage and hypertensive disorders [18].

Comparison of our results with other studies is limited by differences in the case definition criteria used, study location (low- or high-income countries), study population (population-based or hospital-based), and obstetric profile (low- or high-risk). The frequency of SMM cases found in our study was lower than that reported by the Severe Maternal Morbidity Surveillance Network, a nationwide, hospital-based study that also used WHO criteria and estimated a prevalence of women with maternal morbidity of 11.6% [20]. The network includes 27 referral maternity hospitals for high-risk pregnancies in the country, and it is possible that the greater severity of women admitted to these services results in a higher frequency of SMM cases. The SIH/SUS and the ANS database contain information on all hospital inpatient services in the country, including smaller and less complex services.

Another hypothesis is the quality of the records in these systems. A validation study of the SIH/SUS, which used data from a study that collected medical record data as a reference standard, identified low negative likelihood values in the SIH/SUS for several diagnoses, including hypertensive and haemorrhagic complications, which are the main causes of maternal death in the country [21]. No validation studies of the ANS database were identified, but a study that compared the frequency of hospitalizations for abortion using diagnosis and procedure data with diagnosis-only data estimated a 2-fold increase in the number of hospitalizations for abortion [22], demonstrating the underreporting of diagnoses in this database, given that ICD registration is not mandatory.

The profile of SMM cases reflects some similarities in the two databases, such as the high frequency of the severe hypertension criterion, compatible with the pattern of maternal mortality observed in the country, with hypertensive causes being the main cause of maternal death, and the postpartum length of stay greater than seven days, reflecting the longer hospitalization time in women with severe morbidity. However, differences consistent with the characteristics of each sector were also observed. The higher frequency of the criteria “respiratory failure,” “central venous access,” “ICU admission,” and “hysterectomy” in the ANS database, when compared to the SIH/SUS, likely reflects the greater availability of resources in private services. It may also be due to the predominance of procedure-based criteria in this sector, due to low ICD reporting. It is noteworthy that in the private sector, the respiratory failure criterion was measured by the procedure “blood gas analysis,” a procedure not available in the SIH/SUS. The four to six times higher frequency of the criteria “ruptured uterus,” “eclampsia,” “severe preeclampsia,” “abruptio placentae,” and “endometritis” suggests a higher occurrence of complications associated with social vulnerability, a pattern also observed in the analysis of prematurity in publicly and privately funded hospitalizations in Brazil [23].

In both sectors, the highest risk of death was observed for the most severe criteria, particularly shock, central venous access, pulmonary oedema, and hypertensive encephalopathy. The much higher odds ratios for hysterectomy, postpartum hemorrhage, sepse, and shock in the SIH/SUS may reflect greater severity of cases and/or delays in access to appropriate and timely treatment in the public sector, resulting in a higher risk of death. Specifically for the ICU admission criterion, the lower odds ratio of death in the private sector when compared to the public sector may reflect greater access to hospitalization in private services, with less severe conditions among women admitted.

Variable risk of death according to the MNM criterion and higher risk of death in women with a higher number of criteria was also observed in a study conducted by the Maternal Morbidity Surveillance Network that evaluate the WHO MNM criteria [20]. In the predictive model developed, hysterectomy and severe pre-eclampsia presented negative coefficients, which the authors attribute to the fact that severe pre-eclampsia tends to be a transient complication, with effective management strategies resulting in a reduced risk of death, in the same way that hysterectomy is a decisive intervention in the outcome of women with hemorrhage and uterine infection. Both indicate the importance of timely diagnosis and treatment for preventing maternal death. Accordingly, in this study, pre-eclampsia had an odds ratio of less than 1 in the private sector and the high odds ratio observed for the hysterectomy criterion, especially in the public sector, indicates potential barriers to accessing this intervention, with hysterectomy likely being performed as a last resort. Another criterion worth highlighting is blood transfusion, which is reported to increase the frequency of SMM when included in the case definition, especially when the transfused amount is not specified [24]. In this study, blood transfusion presented a high odds ratio of death, demonstrating that it is an important marker of SMM in the Brazilian context.

A higher proportion of deaths was observed in the ANS database, both for total hospitalizations and for women with up to four SMM criteria. There are several possible explanations for this result. On the one hand, the higher mortality may reflect the profile of women treated in the private sector, which is composed of older women, with more comorbidities and a higher proportion of cesarean sections, known risk factors for SMM [25] and maternal deaths [26].

However, recording issues in both databases may have contributed to these results. It’s possible that SMM cases in the ANS database are not being identified because ICD registration is not mandatory, resulting in misclassification. Recording procedures in nonspecific groups (e.g., the “exams” procedure group) or using codes adopted by providers but not standardized by the ANS may also result in less identification of procedure-based criteria. These misclassification errors could explain the higher frequency of deaths in women without SMM criteria, or even according to the number of criteria, if the number of criteria is underestimated.

It is also possible that the number of obstetric hospitalizations may be underestimated in the ANS database. In the SIH/SUS, we used validated criteria based on ICD and procedures to identify obstetric hospitalizations [21]. In the ANS database, the available variable “type of obstetric hospitalization” was used, along with ICD and procedures, but these criteria were not validated, making it possible that the number of obstetric hospitalizations is underestimated. Considering only hospitalizations involving delivery procedures, the estimated proportion of live births financed by supplemental health care in Brazil is 12.6% (data not shown in the table). This figure is much lower than the estimated coverage of women of childbearing age with health insurance plans in Brazil, which is 20%. The ANS has several models for financing hospitalizations. Some hospitals operate with predefined fees for admissions for uncomplicated vaginal births and cesarean sections. If these uncomplicated admissions were not captured by our criteria, there may have been a selection bias, with more admissions with complications being captured in the ANS. In the SIH/SUS, admissions are paid based on the procedures requested, and a lower capture of uncomplicated admissions is not expected. In a study evaluating birth registration coverage in the SIH/SUS, birth registration coverage in the SIH/SUS is growing nationwide, reaching over 80% in 2020 [27].

Failures in recording deaths may also have occurred in both databases. A study that evaluated the coverage of death registration for women of childbearing age in the SIH/SUS, compared to registration in the Mortality Information System, estimated an overall coverage of 78% [28], but with poorer quality of registration in smaller services and those with emergency services. Studies evaluating the quality of death registration in the ANS database were not identified. However, we identified the use of hospital discharge codes in the ANS database that have been disused in the country for many years, which may have resulted in misclassification of the death outcome due to the use of an incorrect code.

Finally, the characteristics of SMM cases in each database may have affected overall mortality. In the SIH/SUS database, more than half of the cases were identified using the hypertensive disorders criterion, which included ICDs corresponding to less severe pathologies, resulting in the lowest odds ratio of death among the four criterion groups. The inclusion of these ICDs in both databases aimed to increase sensitivity in capturing cases of hypertensive disorders, the leading cause of maternal death in Brazil, given that using only ICDs of greater severity could underestimate the importance of hypertensive disorders as a cause of maternal morbidity. On the other hand, in the ANS database, 62.66% of cases were identified using severe management criteria, which are markers of greater severity. Thus, although SIH/SUS presented a higher frequency of SMM cases and a higher OR of death for all criteria, the composition of cases affected overall mortality, resulting in a higher proportion of deaths in the ANS.

This study used administrative databases and is subject to limitations related to coverage and gaps in the recording of each system, as described previously. Another limitation was the use of an algorithm in the SIH/SUS to identify women with care episodes containing more than one AIH [15], which was not possible in the ANS database. Although the ANS guideline recommends using only one identification (ID) during hospitalization, it is possible for the same woman to have more than one ID if there was a change in provider or if a new account was opened without mentioning the previous ID. In the ANS database, 2.9% of hospitalizations were identified with the discharge reason “hospital permanence” and 1.9% with “administrative discharge,” indicating that these women remained hospitalized, likely with a new ID issued. However, unlike the SIH/SUS, it was not possible to identify subsequent hospitalizations during the same hospital stay, as the ANS database does not contain the number of the hospital where the patient was admitted, nor the woman’s date of birth, two essential pieces of information for compiling a care episode with more than one admission form. In the SIH/SUS database, the use of the algorithm resulted in the identification of episodes with longer length of stay, higher hospitalization costs, a higher proportion of ICU admissions, and a higher frequency of deaths [15]. The impossibility of using the algorithm in the ANS database may have resulted in underreporting of complications and deaths.

## Conclusion

Despite the limitations identified in the two administrative databases, this study showed that it is possible to estimate the number of SMM cases in the country, both in hospitalizations with public and private funding, with the results being coherent and compatible with the profile of women hospitalized in both sectors, highlighting the greater probability of death in more severe criteria and in women with a greater number of criteria.

It should be noted, however, that although the same WHO criteria [6] were used to define SMM cases in both databases, their operationalization was different, with more cases based on the severe management criteria in the ANS database. Given these operational differences, adding the cases identified in the two databases to estimate the ratio of SMM to the total number of women would not be appropriate, as this calculation would be affected by the proportion of women treated in the public and private sectors in each location. However, calculating SMM rates for total hospitalizations and monitoring their frequency and key criteria in each sector can provide important insights for planning health interventions.

SMM estimates using the presented methodology have already been calculated for various geographic regions, with the smallest unit of analysis being the 5,570 Brazilian municipalities. The data are available for consultation on the maternal health surveillance dashboard (https://observatorioobstetrico.shinyapps.io/painel-vigilancia-saude-materna/). These data can be used by health surveillance professionals and municipal and state administrations as a strategic tool for formulating policies and actions to reduce maternal mortality in the country and for planning maternal health care services. However, it is important to emphasize the need for continued investment in the quality of recording in both databases so that the estimates can truly reflect the maternal morbidity of Brazilian women and adequately guide strategies to improve the quality of obstetric care in the country.

## Data Availability

All relevant data are within the manuscript. The codes used in the analysis are avaiable at https://www.synapse.org/Synapse:syn64313527/datasets/

## Notes

### Competing Interest Statement

The authors have declared no competing interest.

### Funding Statement

Yes

### Author Declarations

We only used de-identified publicly available data. According to The Brazilian National Health Council Ethics Resolution n? 510/2016 (April 7, 2016) the research ethics committee approval is waived.

